# Hospitalisation for COVID-19 predicts long lasting cerebrovascular impairment: A prospective observational cohort study

**DOI:** 10.1101/2022.02.01.22270235

**Authors:** Kamen A Tsvetanov, Lennart R B Spindler, Emmanuel A Stamatakis, Virginia FJ Newcombe, Victoria C Lupson, Doris A Chatfield, Anne E Manktelow, Joanne G Outtrim, Anne Elmer, Nathalie Kingston, John R Bradley, Edward T Bullmore, James B Rowe, David K Menon, The Cambridge NeuroCOVID Group, The NIHR COVID-19 BioResource, The Cambridge NIHR Clinical Research Facility, The CITIID-NIHR BioResource COVID-19 Collaboration

**Author notes:** Equal contributions (Edward T Bullmore, James B Rowe, David K Menon).

## Abstract

Human coronavirus disease 2019 (COVID-19) due to severe acute respiratory syndrome coronavirus-2 (SARS-CoV-2) has multiple neurological consequences, but its long-term effect on brain health is still uncertain. The cerebrovascular consequences of COVID-19 may also affect brain health. Here we assess cerebrovascular health in 45 hospitalised patients using the resting state fluctuation amplitudes (RSFA) from functional magnetic resonance imaging, in relation to disease severity and in contrast with 42 controls. Widespread changes in frontoparietal RSFA were related to the severity of the acute COVID-19 episode, as indexed by COVID-19 WHO Progression Scale, inflammatory and coagulatory biomarkers. This relationship was not explained by chronic cardiorespiratory dysfunction, age, or sex. Exploratory analysis suggests that the level of cerebrovascular dysfunction is associated with cognitive, mental, and physical health at follow-up. The principal findings were consistent across univariate and multivariate approaches. The results indicate chronic cerebrovascular impairment following severe acute COVID-19, with the potential for long-term consequences on cognitive function and mental wellbeing.

## 1. Introduction

Severe acute respiratory syndrome coronavirus-2 (SARS-CoV-2) causes human coronavirus disease 2019 (COVID-19) with multi-system effects that include neurological, vascular and neurovascular injury. Acute neurological sequelae are common, ranging from mild dizziness, headaches and anosmia to severe encephalitis, stroke and delirium (Chen et al., 2020; Hensley et al., 2021; Paterson et al., 2020; Zubair et al., 2020). These sequelae may arise from systemic physiological insults (e.g. hypoxia, hypotension, dysautonomia), coagulation dysfunction, large vessel occlusion, arterial stiffness, impaired vasoreactivity, neurotropic infection, parenchymal haemorrhage, or autoimmune responses against diverse antigens (Chen et al., 2020; Marcic et al., 2021; Mohkhedkar et al., 2021; Schnaubelt et al., 2021). Acute COVID-19 has also been associated with microvascular injury from vasculitis or endothelialitis (Becker, 2020; McGonagle et al., 2021), with endotheliopathy (Kakarla et al., 2021), vasogenic oedema and microthrombosis in the acute phase (Iba et al., 2020; Levi et al., 2020) and hypoperfusion in subacute phase (Hosp et al., 2021). While this acute pathophysiology is detectable using neuroimaging (Hanafi et al., 2020; Lersy et al., 2021; Newcombe et al., 2020), the persistence and effects of cerebrovascular dysfunction over the medium- and long-term remain unknown.

An important aspect of cerebrovascular function is the capacity of cerebral vessels to constrict or dilate in response to physiological conditions such as alterations in carbon dioxide (CO2) and oxygen tension. This cerebrovascular reactivity (Willie et al., 2014) regulates regional blood flow via pH-dependent modulation of vascular smooth muscle tone (Ainslie et al., 2005; Jensen et al., 1988; Lambertsen et al., 1961; Lassen, 1968), but is compromised by arterial stiffness, compromised endothelial function (Brandes et al., 2005), or disorders including hypertension, traumatic brain injury and dementia. Poor cerebrovascular reactivity may also increase the risk of neurodegeneration (Gao et al., 2013).

We therefore assessed the impact of COVID-19 on chronic cerebrovascular reactivity after hospitalisation. We use a well-established non-invasive imaging method, exploiting naturally occurring fluctuations in arterial CO2 induced by variations in cardiac and respiratory cycles, which moderate the blood oxygenation level-dependent (BOLD) signal underlying functional magnetic resonance imaging (Birn et al., 2006; Glover et al., 2007). The BOLD signal variability at rest, known as resting state fluctuation amplitudes (RSFA) is a safe, scalable and robust alternative to the gold standard approaches (Kannurpatti and Biswal, 2008; Tsvetanov et al., 2020c, 2015). It is easier and safer to apply in clinical cohorts than experimental hypercapnia, breath-holding and drug interventions (Keyeux et al., 1995; Rostrup et al., 1996, 1994; Wagerle and Mishra, 1988). RSFA is sensitive to cerebrovascular and cardiovascular differences in ageing (Garrett et al., 2017; Tsvetanov et al., 2020b, 2015), small vessel disease (Makedonov et al., 2013), stroke (Nair et al., 2017; Raut et al., 2016), Alzheimer’s disease (Makedonov et al., 2016; Millar et al., 2020a), cognitive performance (Liu et al., 2021; Millar et al., 2021, 2020b) and the presence of brain tumours (Agarwal et al., 2019, 2017).

We report on the chronic effect of COVID-19 on RSFA as a marker of cerebral microvascular function, in relation to acute severity. Exploratory analyses examine the relationship to normative regional variations in neurotransmitters/receptors, brain energy consumption and cell-type distributions. Acute disease severity was quantified by the COVID-19 WHO Progression Scale and blood biomarkers in patients hospitalised for COVID-19. We predicted that acute COVID-19 changes regional RSFA at follow up, in proportion to acute disease severity, over and above the effects of residual systemic cardiorespiratory impairment.

## 2. Methods

### 2.1. Participants

Patients were recruited through the NIHR COVID-19 BioResource, which received ethical approval from East of England - Cambridge Central Research Ethics Committee (REC 17/EE/0025), and provided written informed consent. Eligibility was based on admission to Addenbrookes Hospital with COVID-19 between 10th March 2020 and 31st July 2020, aged 18 years or older, survived the acute illness, and attended for a follow up visit, and no contradictions to MRI. 489 patients were potentially eligible. Clinical data were obtained from inpatient electronic medical records, and from cardiorespiratory and neurological assessment at follow-up clinical and research visits at least 6 weeks following symptom onset. 45 patients consented to participate, with clinical, structural and functional resting state functional magnetic resonance imaging (fMRI) data of appropriate quality (see below). Age and sex matched controls were scanned with the same sequences, with data pooled over sequential cohorts to match demographics (Cambridgeshire Research Ethics Committee 97/290 and REC 17/EE/0025, and Norfolk EE/0395 and a protocol approved by the Human Biology Ethics Committee of the Council of the School of Biological Sciences, University of Cambridge). The study and their processing pipelines are summarised in SI Figure 1.

**Figure 1.**
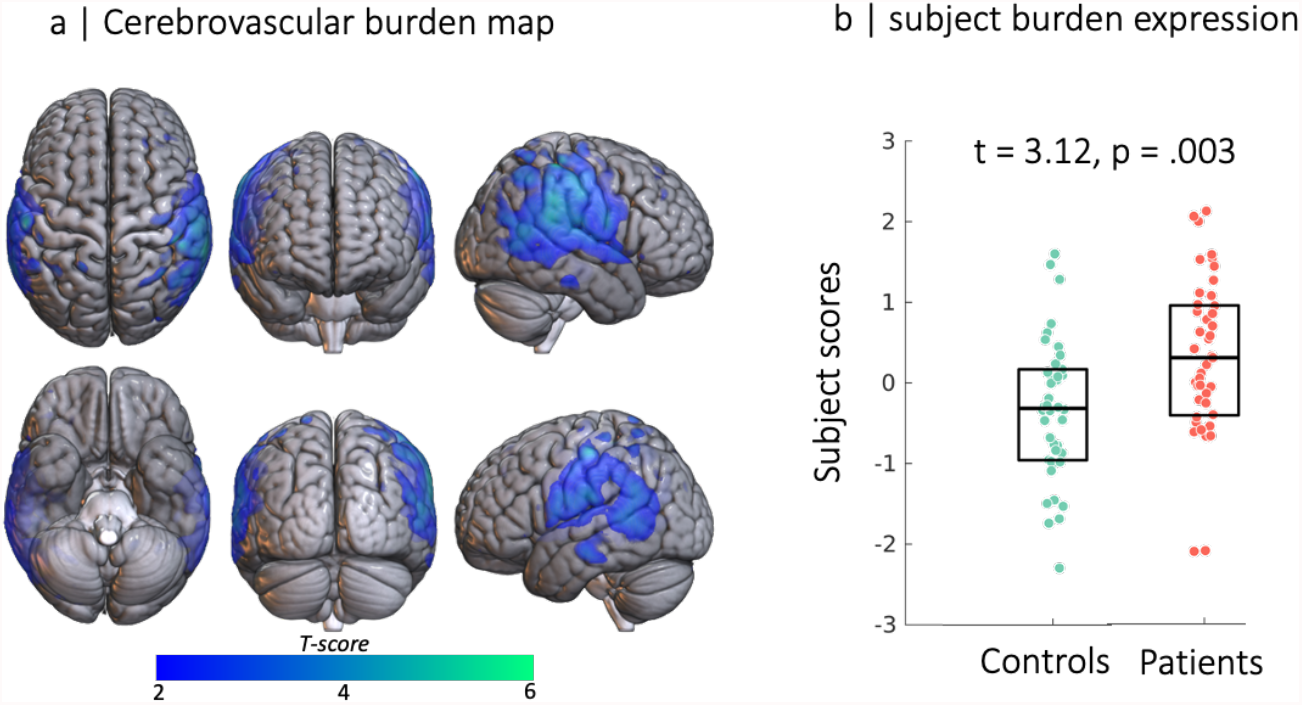
Source-based cerebrovasculometry for the component differentially expressed between groups: (left) independent component spatial map reflecting decrease in RSFA values in temporo-parietal regions. (right) Bar plot of subject scores for patients hospitalised for COVID-19 (red) and control group (green, each circle represents an individual) indicating higher loading values for patients than controls as informed by two-sample unpaired permutation test (a robust regression was used to down-weight the effects of extreme data points)

### 2.2. Inpatient data: COVID-19 severity

The WHO COVID-19 11-point Progression scale was used to provide a measure of disease severity with scores from 0 (non-infected) to 10 (dead) (see Figure 1 in Marshall et al., 2020). We also used blood biomarkers previously associated with COVID-19 severity including the most extreme values during hospitalisation on haematological (lowest platelets) (Wool and Miller, 2021), inflammatory (C-reactive protein, CRP, serum ferritin) (Luan et al., 2021), immunological (inteleukin-6, IL-6) (Group et al., 2021a; Ulhaq and Soraya, 2020), hepatic (bilirubin) (Bangash et al., 2020) and coagulatory (D-dimer, prothrombin time (PT) and activated partial thromboplastin time (APTT)) (Asakura and Ogawa, 2020; Iba et al., 2020; Levi et al., 2020). Details of laboratory assay methods used are provided in the Supplementary section. All blood-based measurements were log-transformed to transform the skewed data to approximately conform to normality. For consistency in interpreting scores across blood samples, platelets values were inverted (iPlatelets) so that higher scores represent lower count.

### 2.3. Clinical visit: cardiorespiratory assessment

Cardiorespiratory measurements were collected during a clinical assessment at least 12 weeks after discharge from initial hospitalization with COVID-19. Systolic and diastolic blood pressure (BPS and BPD) were measured in lying (or seated) and standing position using automated sphygmomanometry using GE Carescape V100 Dinamap Vital Signs Monitor (GE Healthcare Systems, Chicago, Illinois, USA). We calculated pulse pressure (BPS-BPD) and orthostatic hypotension (e.g. BPS lying – BPS standing). Lying and standing pulse pressure values were log-transformed to transform the skewed data to approximately conform to normality.

Lung function was assessed via CareFusion Micro 1 Handheld Spirometer (Vyairre Medical GmbH.; Hoechenberg, Germany) to determine peak expiratory flow (PEF), forced expiratory volume in 1 second (FEV1), forced vital capacity (FVC) and FEV1/FVC ratio. Measurements were repeated in triplicate, with one minute rest between measurements. Each variable was log-transformed to approximately conform to normality. To balance the representativeness of each data type in the imputations and data-reduction stages (see Supplementary Method), spirometry measures were reduced to a single variable using principal components analyses (SI Figure 2).

**Figure 2.**
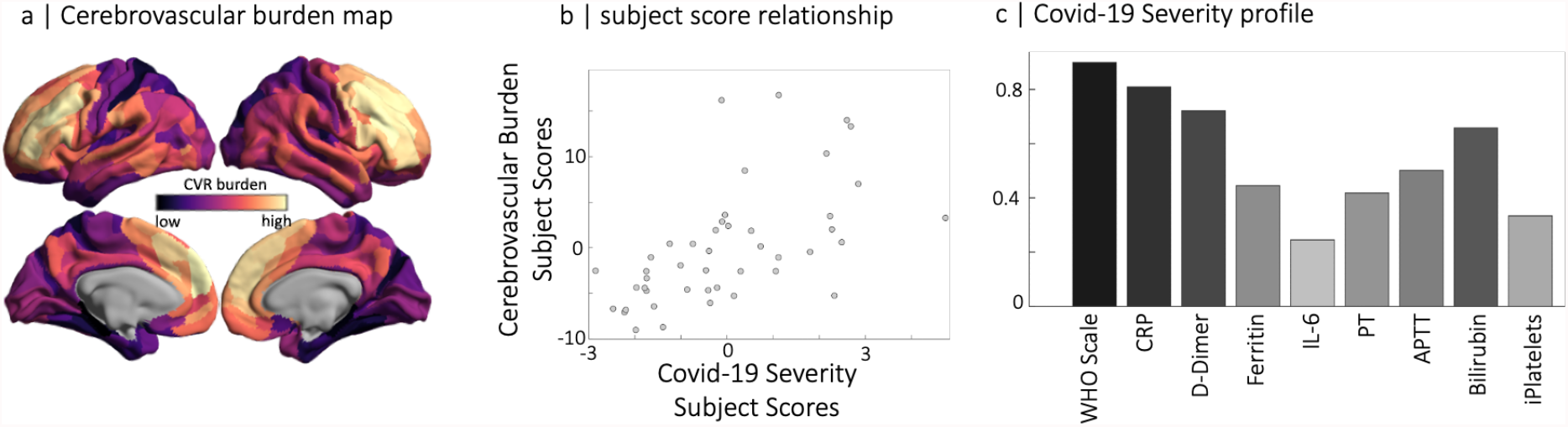
Partial least squares analysis of COVID-19 severity data at acute stage and RSFA-based cerebrovascular burden (CVB) at chronic stage. (panel a) Spatial distribution of parcellated RSFA values where dark to light colours are used for the strength of positive and negative correlations with the COVID-19 Severity profile (panel c). Note that regions with high cerebrovascular burden have low values in RSFA. The scatter plot in the middle panel represents the relationship between subjects scores of RSFA-latent variable and COVID-19 Severity-latent variable identified by partial least squares analysis.

Pulse oximetry, monitored via GE Carescape V100 Dinamap Vital Signs Monitor (GE Healthcare Systems, Chicago, Illinois, USA), was used to determine the heart rate and arterial oxygen saturation before and following a 6-minute walk test (Bois et al., 2012; Crapo et al., 2012; PL et al., 2003).

### 2.4. Research visit: neurological assessment

#### 2.4.1. Image acquisition and pre-processing

Imaging data were acquired using a 3T Siemens Prisma*fit* System with a 32-/20-channel head-coil at the Wolfson Brain Imaging Centre (WBIC; www.wbic.cam.ac.uk) during a research visit. A 3D-structural MRI was acquired on each participant using T1-weighted sequence (3D Magenetisation-Prepared Rapid Gradient-Echo, 3D MPRAGE) with the following parameters: repetition time (TR) = 2 ms; echo time (TE) = 2.99 ms; inversion time (TI) = 880 ms; flip angle α= 9°; field of view (FOV) = 208 × 256 × 256 mm3; resolution = 1 mm isotropic; accelerated factor (in-plane acceleration iPAT) = 2; acquisition time, 5 min. T1 images were pre-processed using functions from SPM12 (Wellcome Department of Imaging Neuroscience, London, UK) in Matlab 2020b (Mathworks, https://uk.mathworks.com/). The T1 image was rigid-body coregistered to the MNI template, and segmented to extract probabilistic maps of six tissue classes: gray matter (GM), white matter (WM), CSF, bone, soft tissue, and residual noise.

RSFA was estimated from resting state Echo-Planar Imaging (EPI) of 477 volumes acquired with 64 slices for whole brain coverage (TR = 735ms; TE = 30ms; FOV = 210mm x 210mm; resolution = 2.38 × 2.38 × 2.4mm) during 5 minutes and 51 seconds. Participants were instructed to lay still, to stay away and keep their eyes open, looking at a fixation cross. EPI data preprocessing included the following steps: (1) temporal realignment of slices to (0,0,0) Montreal Neurological Institute (MNI) co-ordinates, (2) spatial realignment to adjust for linear head motion, (3) subjected to Artifact detection tools (ART)-based identification of outlier scans for scrubbing (4) coregistration to the T1 anatomical image from above, (4) application of the normalization parameters from the T1 stream above to warp the functional images into MNI space. We applied whole-brain independent component analysis of sing-subject time series denoising to minimise motion artefacts using a priori heristics using the ICA-based Automatic Removal of Motion Artifact toolbox (AROMA; Pruim et al., 2015a, 2015b) after smoothing with an 6mm FWHM Gaussian kernel.

The initial six volumes were discarded to allow for T1 equilibration. We quantified participant motion using the root mean square volume-to-volume displacement as per Jenkinson et al (2002). A general linear model (GLM) of the time-course of each voxel was used to further reduce the effects of noise confounds (Geerligs et al., 2017), with linear trends and expansions of realignment parameters, plus average signal in WM and CSF, CompCorr regressors, their derivative and quadratic regressors (Satterthwaite et al., 2013). The WM and CSF signal was created by using the average across all voxels with corresponding tissue probability larger than 0.7 in associated tissue probability maps available in SPM12. A band-pass filter (0.0078-0.1 Hz) was implemented by including a discrete cosine transform set in the GLM, ensuring that nuisance regression and filtering were performed simultaneously (Hallquist et al., 2013; Lindquist et al., 2019). Finally, we calculated subject specific maps of RSFA based on the normalized standard deviation across time for processed resting state fMRI time series data (Tsvetanov et al., 2020b). RSFA maps were smoothed by a 12 mm FWHM Gaussian kernel.

To facilitate integrative multivariate analysis (see below), the RSFA maps were parcellated by a prior cortical template into 360 bilaterally symmetric regions (Glasser et al., 2016). Regional RSFA values were estimated by averaging over all voxels in each parcel.

#### 2.4.2. Quality of life, cognition, and mental health

Quality of life, cognition and mental health were assessed using a set of questionnaires: Generalised Anxiety Disorder-7 (GAD-7), Patient Health Questionnaire-9 (PHQ-9), Patient Health Questionnaire-15 (PHQ-15), Posttraumatic Stress Disorder Checklist-5 (PCL-5) and subscores from the Short Form-36 (SF-36). SF36 subscores were defined as physical functioning (SF36-PF), role limitation physical (SF36-RLP), role limitation emotional (SF36-RLE), energy dimension (SF36-ED), emotional wellbeing (SF36-EW), social functioning (SF36-SF), pain (SF36-P) and general health (SF36-GH).

Cognitive function and functional independence were evaluated using Montreal Cognitive Assessment (MoCA, Nasreddine et al., 2005), inverted Modified Ranking Scale (mRS, Eriksson et al., 2007), and Barthel Index (BI, Mahoney and Barthel, 1965). For consistency in interpreting scores across questionnaires, scores for mental health questionnaires were inverted (iGAD-7, iPHQ-9, iPHQ-15 and iPCL-5) so that lower values represent greater mental health problems.

### 2.5. Analytical approach

Statistical analysis used Matlab 2020b calling the packages as described below. We performed a descriptive analysis of all the characteristics from inpatient, clinical and cognitive data before integrating it with neuroimaging data as described below.

#### 2.5.1. Group differences in cerebral microvascular health

We performed component-based analysis, Source-Based Cerebrovasculometry (Supplementary Material) to determine spatially non-overlapping RSFA maps without using group information. Subject scores for each component was predicted by group identity (Patient vs Control), age, and sex in a subsequence robust multiple linear regression. The model’s formula was specified by Wilkinson’s notation, ‘rsfa ∼ group*age + sex’ and fitted for each component separately.

#### 2.5.2. COVID-19 severity predicting RSFA

The predictive specificity of COVID-19 severity on RSFA abnormality was tested using a multivariate approach. We adopted a two-level procedure (Passamonti et al., 2019a; Tsvetanov et al., 2020a, 2018, 2016). In the first-level analysis, the relationships between COVID-19 Severity and RSFA data were identified using partial least squares (Krishnan et al., 2011). Partial least squares described the linear relationship between the two multivariate data sets, namely RSFA maps and COVID-19 Severity data by providing pairs of latent variables (RSFA-LV) and (COVID-19 Severity-LV) as linear combinations of the original variables that were optimized to maximize their covariance. Data set 1 consisted of parcellated RSFA maps across all patients (45 patients x 360 nodes array, RSFA dataset). Data set 2 included the COVID-19 WHO Progression scale and inpatient blood data (45 subjects x 9 measures array, COVID-19 Severity datasets). All variables were z-scored (mean of 0 and standard deviation of 1) before entering to a permutation-based PLS analysis with 10.000 permutations to determine the significance of the latent variables.

Next, we tested whether the identified relationship between COVID-19 Severity-LV and RSFA-LV could be explained by other variables of interest and variables of no interest. To this end, we performed a second-level analysis using robust multiple linear regression and commonality analysis (Kraha et al., 2012; Nimon et al., 2008a). Commonality analysis partitions the variance explained by all predictors in MLR into variance unique to each predictor and variance shared between each combination of predictors. Therefore, unique effects indicate the (orthogonal) variance explained by one predictor over and above that explained by other predictors in the model, while common effects indicate the variance shared between correlated predictors. Notably, the sum of variances, also known as commonality coefficients, equals the total R2 for the regression model. We adapted a commonality analysis algorithm (Nimon et al., 2008b) implemented in Matlab (Wu et al., 2021).This model aimed to test whether the relationship between COVID-19 Severity and RSFA can be explained partly or fully by systemic cardiorespiratory dysfunction or other covariates of no interest. Predictor variables included subject COVID-19 Severity scores from the first level PLS and systemic cardiorespiratory dysfunction (CRDPC1 and CRDPC2). The dependent variable was subjects’ RSFA scores from the first level PLS. Covariates of no interest included age and sex. The model can therefore identify unique variance explained by each of the predictors i.e. whether COVID-19 Severity LV predicts RSFA-LV over and above other predictors. Common effects of interest were the cardiorespiratory-related effects, defined by the common variance between COVID-19 Severity and CRDPCA1-2. Significant effects of interest were identified with nonparametric testing using 10.000 permutations using commonality analysis implementation in Matlab (Wu et al., 2021).

#### 2.5.3. Exploratory analysis

In an exploratory analysis, the level of RSFA abnormalities was related to component measures of physical, cognitive, and mental functioning (PCM) using robust regression. The PCM components, defined by principal component analysis (SI Methods), we defined as dependent variables in separate models. RSFA, age and sex were entered as predictors. The model’s formula was specified by Wilkinson’s notation, ‘PCM ∼ RSFA + age + sex’ and fitted for each PCM component separately.

We further assessed the spatial overlap between COVID-19-related cerebrovascular burden maps and a range of brain metabolic, neurotransmitter, protein expression and cell-type parameters, including i) existing receptor/metabolic templates and ii) gene transcription profiling maps. Templates of interest included metabolic rates of glucose, oxygen, and aerobic glycolysis (Vaishnavi et al., 2010) and receptor and transmitter maps across nine different neurotransmitter systems (Hansen et al., 2021b), all measured by positron emission tomography (PET). Gene expression maps (Hawrylycz et al., 2012) were based on key proteins implicated in SARS-Cov-2 cellular attachment (angiotensin converting enzyme-2, ACE2; neuropilin-1, NRP1; neuropilin-2, NRP2), processing (cathepsin-B, CTSB; cathepsin-L, CTSL) and viral defence (interferon type 2 receptors, IFNAR2; lymphocyte antigen 6-family member E, LY6E) (Iadecola et al., 2020; Yang et al., 2021). Spatial correlations were evaluated using spin-based permutations preserving spatial autocorrelation (spin-permutation test) (Alexander-Bloch et al., 2018; Fulcher et al., 2021).

Spatial covariance between the cerebrovascular burden map and gene expression across 9394 genes expressed in the human brain (Hawrylycz et al., 2012) was based on regularized-PLS association (Blankertz et al., 2011; Ledoit and Wolf, 2004) using spin-premutation-based 10-fold cross-validations (Tsvetanov et al., 2020a). The latent variable represented a spatial pattern of gene expression that highly covaries with the cerebrovascular burden pattern associated with COVID-19 severity. Full details about the processed transcriptomic data are available elsewhere (Arnatkeviciūtė et al., 2019). Genes highly expressing this pattern (i.e. high loadings) were tested against a molecular atlas of human brain vasculature of 17 control and Alzheimer’s disease patients (Yang et al., 2021) using cell-type decomposition approach (Hansen et al., 2021a). This enabled us to test whether the cerebrovascular burden-relevant genes are preferentially expressing in specific cell types i.e. testing for cell-specific aggregate gene sets in eleven major canonical cortical cell classes: astrocytes (Astro); brain endothelial cells (BEC); ependymal (Epend); macrophage/microglia (MacMic); meningeal fibroblast (MFibro); neuron (Neuro); oligodendrocyte precursors (Opc); oligodendrocytes (Oligo); pericytes (Peri); perivascular fibroblast (PFibro); smooth muscle cells (SMC). To this end, we calculated the ratio of genes in each set preferentially expressed in each cell type (e.g. ratio for pericytes is calculated from the number of genes preferentially expressed in pericytes divided by the total number of genes). Gene sets were thresholded to include the top *n*% of genes with greatest loadings, where *n* varied from 5% to no threshold. Statistical significance was determined using a null distribution of ratios based on 10.000 sets of random genes (Hansen et al., 2021a).

### 2.6. Data availability and code

Code and composite data to reproduce manuscript figures, statistical analyses are available at https://github.com/kamentsvetanov/covid19_cerebrovascularburden. Resting-state fMRI data was pre-processed using SPM12 (http://www.fil.ion.ucl.ac.uk/spm; Friston et al., 2007) and post-processed using a GLM-like approach (Geerligs et al., 2017) available at https://github.com/MRC-CBU/riksneurotools/blob/master/GLM/. MATLAB-based commonality analysis for neuroimaging is available at https://github.com/kamentsvetanov/CommonalityAnalysis/. Visualisation of neuroimaging results was in MRIcroGL (https://github.com/rordenlab/MRIcroGL; Rorden and Brett, 2000) and BrainSpace (Vos de Wael et al., 2020). Neurotransmitter receptor and transporter maps were available at https://github.com/netneurolab/hansen_receptors. Spin permutations (Váša et al., 2018) used code available at https://github.com/frantisekvasa/rotate_parcellation. Fully-pre-processed transcriptomic data was available at https://figshare.com/articles/dataset/AHBAdata/6852911 and https://github.com/BMHLab/AHBAprocessing. The molecular atlas of the human brain vasculature was available at https://www.biorxiv.org/content/10.1101/2021.04.26.441262v1. Cell-type decomposition analysis related code was available at https://github.com/netneurolab/hansen_genescognition.

## 3. Results

### 3.1. Participants

Characteristics of the 45 patients hospitalized with COVID-19 are detailed in Table 1. At hospitalisation, 30% of the patients required a mechanical ventilation (n=13) for an average of 20 days; 22% of patients required vasopressors (n=10), and 9% renal replacement therapy with continuous veno-venous haemodiafiltration (n=4). The average number of days from initial symptoms to clinical and research visit was 169±35 and 180±58, respectively. Data missingness is detailed in Supplementary Results.

**Table 1.** Characteristics of 45 patients hospitalised with COVID-19. n - indicates number of patients and the percentage of patients from the patient cohort (%), SD – standard deviation, IQR – interquartile range, kg – weight in kilograms, m – height in meters.

| Variable | n (%) | Mean(SD)/Median(IQR)* |
| --- | --- | --- |
| Age (years) | 45 (100) | 52 (14) |
| Education (years) | 45 (100) | 16 (4) |
| Female | 26 (58) | - |
| Right-handed | 40 (89) | - |
| Inpatient data |  |  |
| Vasopressors | 10 (22) | - |
| Dialysis | 4 (9) | - |
| BMI (kg/m<sup>2</sup>) | 36 (80) | 29 (4) |
| Mechanical Ventilation (days) | 13 (29) | 20 (15) |
| CRP | 42 (93) | 149 (147) |
| D Dimer | 37 (82) | 1125 (2071) |
| Ferritin | 32 (71) | 1225 (1700) |
| IL-6 | 28 (62) | 39 (80) |
| PT | 32 (71) | 26 (54) |
| APTT | 33 (73) | 42 (36) |
| Bilirubin | 43 (96) | 14 (8) |
| Platelets | 44 (98) | 230 (102) |
| Clinical Visit |  |  |
| HR (bpm) | 38 (84) | 76 (14) |
| RR (cpm) | 38 (84) | 16 (2) |
| SBP lying (mmHg) | 38 (84) | 134 (23) |
| SBP standing (mmHg) | 33 (73) | 130 (20) |
| DBP lying (mmHg) | 38 (84) | 74 (12) |
| DBP standing (mmHg) | 33 (73) | 79 (11) |
| Research Visit |  |  |
| GAD-7 (0-21) | 44 (98) | 4 (8)* |
| PHQ-9 (0-27) | 44 (98) | 7 (9)* |
| PHHQ-15 (0-30) | 39 (87) | 9 (8)* |
| PCL-5 (0-80) | 44 (98) | 16 (22)* |
| MoCA | 39 (87) | 28 (3)* |
| mRS | 39 (87) | 1 (1)* |
| BI | 37 (82) | 20 (0)* |
| Initial symptoms to Admission (days) | 40 (89) | 14 (14) |
| Hospitalisation Duration (days) | 40 (89) | 19 (31) |
| Initial symptoms to Clinic visit (days) | 39 (87) | 169 (35) |
| Initial symptoms to Research visit (days) | 45 (100) | 180 (58) |

### 3.2. Group differences in Cerebrovascular components

Using source-based cerebrovasculometry (Passamonti et al., 2019b; Tsvetanov et al., 2020b, 2015; Xu et al., 2009), we assessed the group differences in RSFA-based cerebrovascular components between controls and patients in small set of spatially independent components (n=8). Each participant had one value per component, *subject score*, indicating the degree to which a participant expresses the spatial map of the corresponding component. One component showed significant difference between the patient and control group in terms of their subject scores (RSFAIC4, t = 3.12, p = 0.003, Figure 1), while controlling for age and sex in a robust linear regression. The spatial extent of this component, RSFAIC4, included voxels with high values in temporo-parietal regions, indicating that individuals with higher loading values, in this case the patient group, had lower RSFA values in these regions, relative to the control group (Figure 1). The other components did not differentiate patients from controls (SI Analyses).

The spatial pattern of IC4 was consistent with the univariate approach (r=0.46, p<0.001, SI Figure 5). In addition, the univariate approach revealed that the spatial pattern in RSFA associated with age was highly consistent with the one reported on large-scale population-based cohorts, r=0.42, p<0.001 (Tsvetanov et al., 2020b, 2015). This suggests that RSFA can detect reliably differences in cerebrovascular health across various phenotypes in smaller samples. Though age is a risk factor of COVID-19 severity (Verity et al., 2020) and RSFA (Tsvetanov et al., 2020c), the COVID-19 group effect was not explained by individual’s age, and showed only a partial overlap with the effects of age on RSFA in parietal regions (SI Figure 5, Tsvetanov et al., 2020b, 2015)0.

### 3.3. COVID-19 Severity predicts cerebrovascular impairment

Using PLS analysis, we identified one significant pair of latent variables (r = 0.584, p = 0.017, based on a null distribution of 10,000 permutations). Variable loadings and subject scores showed a strong relationship between acute COVID-19 Severity and chronic RSFA abnormalities (Figure 2).

The RSFA latent variable (RSFA-LV) expressed negative loadings in frontal (superior frontal gyrus, middle frontal gyrus, inferior frontal gyrus and portions of the anterior cingulate) and parieto-temporal (angular gyrus, supramarginal gyrus, superior temporal gurus, middle temporal gyrus) regions. This pattern of COVID-19 Severity-related reduction in RSFA values was mirrored in a voxel-wise analysis of RSFA maps and COVID-19 Severity component (SI Figure 5). Positive loadings in the RSFA data appeared to be in postcentral gyrus, calcarine sulcus, cuneus, and lingual gyrus. Increase in the RSFA signal in these regions may reflect increased pulsatility in neighbouring vascular and white matter territories (see SI Figure 5) as reported previously (Makedonov et al., 2016, 2013; Tsvetanov et al., 2020b, 2015). This pair of latent variables suggested that patients with higher COVID-19 Severity at acute stage have poorer cerebrovascular function in frontal and temporo-parietal regions at chronic stage. For visualisation purposes we inverted the loading values in Figure 2a so that higher values reflect poorer cerebrovascular function, i.e. higher cerebrovascular burden.

To understand whether the relationship between regional RSFA impairment and COVID-19 Severity can be explained by chronic systemic cardiorespiratory dysfunction (see SI method) or covariates of no interest, we performed a second level robust regression analysis. RSFA-LV was entered as the dependent variable. COVID-19 Severity LV and the cardiorespiratory dysfunction component (CRDPCA1, see SI Figure 3) were entered as predictors, while age and sex were modelled as covariates of no interest. All variables were normalised to have a mean of 0 and standard deviation 1. Using Wilkinson notation, the regression model took the form: *RSFA-LV ∼ COVID-19 Severity-LV + CRDPCA1 + CRDPCA2 + Age + Sex*. COVID-19 Severity was the only significant predictor of RSFA-LV in the model (r = 0.471, p = 0.001 and SI Table 1), suggesting that chronic cardiorespiratory dysfunction, age and sex cannot explain fully the relationship between COVID-19 Severity and RSFA abnormality. Interestingly, the unique variance explained by COVID-19 Severity in the regression model was weaker than the variance identified by the PLS analysis (r = 0.584 vs r=0.471 for PLS and MLR analyses) suggesting that one or more of the predictors in the model explain partly some of the variance between COVID-19 Severity and RSFA. Permutation-based commonality analysis with 10.000 permutations (SI Method) confirmed that a portion of the variance between COVID-19 Severity-LV and RSFA-LV was explained by age (12% total, p < 0.001), cardiorespiratory dysfunction component 2 (CRDPC2, 3% total, p = 0.009), and shared variance between age and cardiorespiratory dysfunction component 1 (Age,CRDPCA1, 20% total, p < 0.001), SI Table 2. COVID-19 Severity-LV remained as the largest unique predictor or RSFA-LV (37% total, p=0.006).

**Figure 3.**
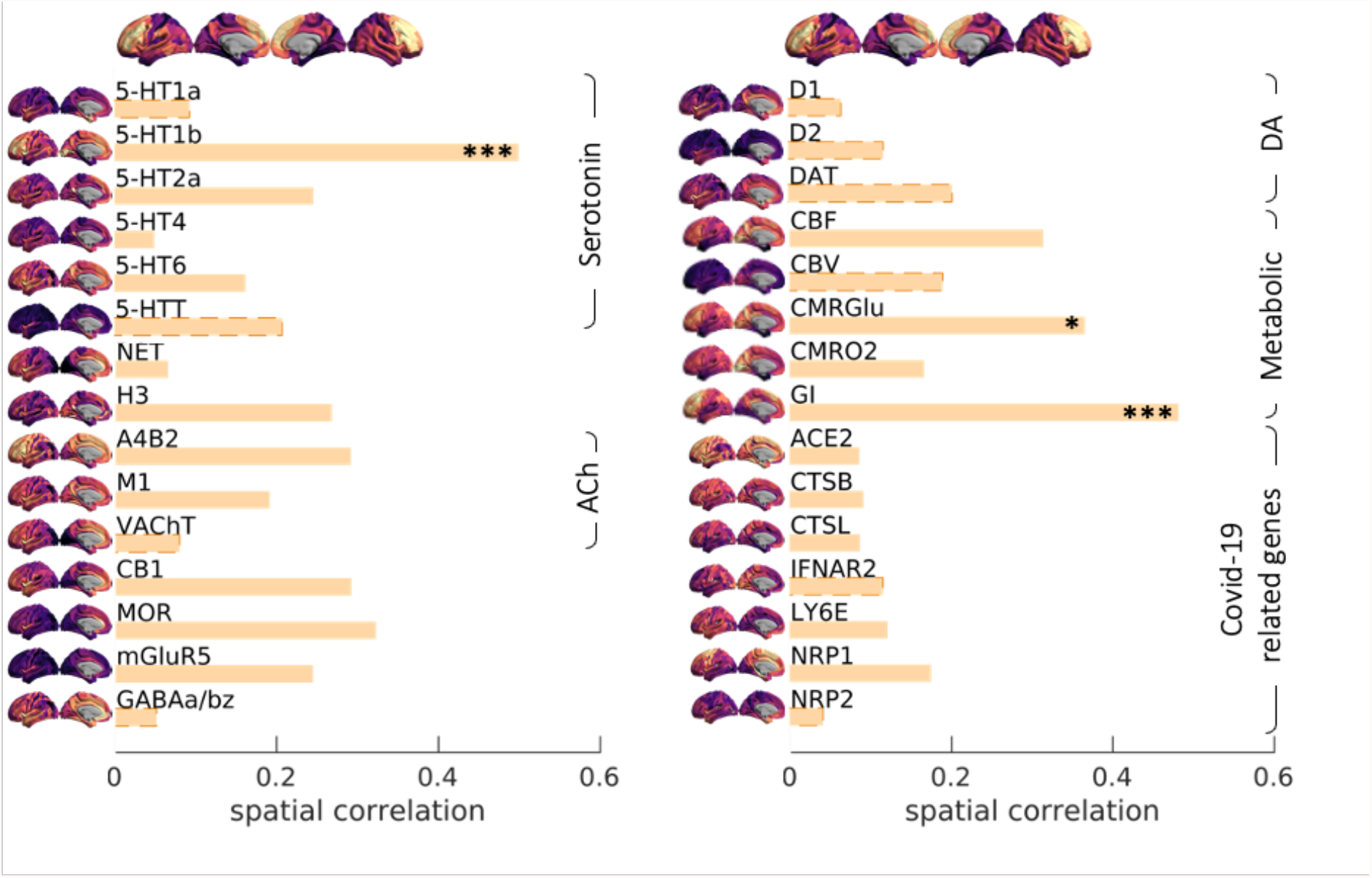
Spatial correlation between Covid19 severity-induced cerebrovascular burden map and spatial patterns associated with a range of neurotransmitter receptor/transporters (Hansen et al., 2021b), selective genes relevant to SARS-CoV-2 brain entry (Iadecola et al., 2020) and brain metabolism parameters (Vaishnavi et al., 2010). Neurotransmitter receptors and transporters were selective to serotonin (5-HT1a, 5-HT1b, 5-HT2a, 5-HT4, 5-HT6, 5-HTT), norepinephrine (NET), histamine (H3), acetylcholine (ACh, A4B2, M1, VAChT), cannabinoid (CB1), opioid (MOR), glutamate (mGluR5), GABA (GABAa/bz) and dopamine (D1, D2, DAT). Metabolic maps were based on cerebal blood flow (CBF), cerebral blood volume (CBV), cerebral metabolic rate of glucose and oxygen (CMRGlu, CMRO2) and glycemic index (GI). Selective genes relevant to SARS-CoV-2 brain entry included angiotensin converting enzyme-2, ACE2; neuropilin-1, NRP1; neuropilin-2, NRP2, cathepsin-B, CTSB; cathepsin-L, CTSL, interferon type 2 receptors, IFNAR2; lymphocyte antigen 6-family member E, LY6E. The spatial maps of 5-HT1b, CMRGlu and Glycemic Index (GI) were significantly correlated with Covid19 severity-induced cerebrovascular burden map (* p-spin<0.05 (one-sided), *** p-spin<0.001). See text for more information.

### 3.4. Exploratory analysis

The level of RSFA abnormalities (RSFA-LV) was related to physical, cognitive and mental functioning in an exploratory analysis. RSFA-LV, age and sex were entered as predictors, while cognitive and mental health components (SI Methods) were used as dependent variables in separate robust regressions (Model 1: CMHPCA1 ∼ RSFA-LV + age + sex; Model 2: CMHPCA2 ∼ RSFA-LV + age + sex). Model 1 was not significant (p = 0.429), while Model 2 was significant (R2 = 0.294, p = 0.002) with RSFA predicting significantly cognitive and mental health component 2 (r = -0.352, p = 0.014, SI Table 3). This indicates that patients with higher RSFA abnormality have worse cognitive function, less functional independence and lower quality of life (social functioning, emotional wellbeing, energy dimension). The results did not change in a meaningful way by re-imputing the data (n=5). The findings in Model 2 were confirmed using voxel-wise analysis on RSFA maps, instead of RSFA-LV (SI Figure 7).

We next assessed the spatial overlap between COVID-19-related cerebrovascular burden maps with existing neurotransmitter and metabolic maps using spatial autocorrelation-preserving permutation testing (Alexander-Bloch et al., 2018). Across 21 candidate maps, we show that the cerebrovascular burden map overlaps with the distribution of serotonin’s vasoactive receptor 5-HT1b, aerobic glycolysis and to a weaker extent cerebral metabolic rate of glucose in the brain. However, the regional distribution of RSFA abnormality showed little correlation with the expression of key proteins implicated in SARS-CoV-2 cellular attachment, processing and viral defence (Figure 3). Collectively, these results demonstrate that the distribution of cerebrovascular impairment related to COVID-19 severity is aligned with the spatial distribution of receptors and processes involved in the coordination of metabolic and vasoreactive responses.

As a final step, we used normative transcriptomics to identify genes that are normally preferentially expressed in the regions associated with COVID-19-induced cerebrovascular impairment. Regularised-PLS identified one latent component (r=.64, p=0.001, Figure 4a). Using cell-type decomposition analysis on a vascular cell-type specific gene sets, we determined the ratio of genes in each gene set preferentially expressed in one of eleven cortical cell types: astrocytes, brain endothelial cells, ependymal, macrophage/microglia, meningeal fibroblast, neuron, oligodendrocyte precursors, oligodendrocytes, pericytes, perivascular fibroblast, smooth muscle cells. Gene-sets were thresholded to include the top 70% of genes with the highest loadings (Figure 4b), noting that results are consistent across thresholds ranging from 5 to no threshold (Supplementary Figure 8). Dominant genes were significantly more expressed in pericytes, brain endothelial cells and neurons, and significantly less expressed in oligodendrocytes. Broadly, we find evidence that areas associated with cerebrovascular impairment are enriched in for genetic signal related to neuron support (pericytes, endothelial cells and perineuronal oligodendrocytes) and neurons themselves. This dichotomy is consistent with the observations from the spatial overlap analysis.

**Figure 4.**
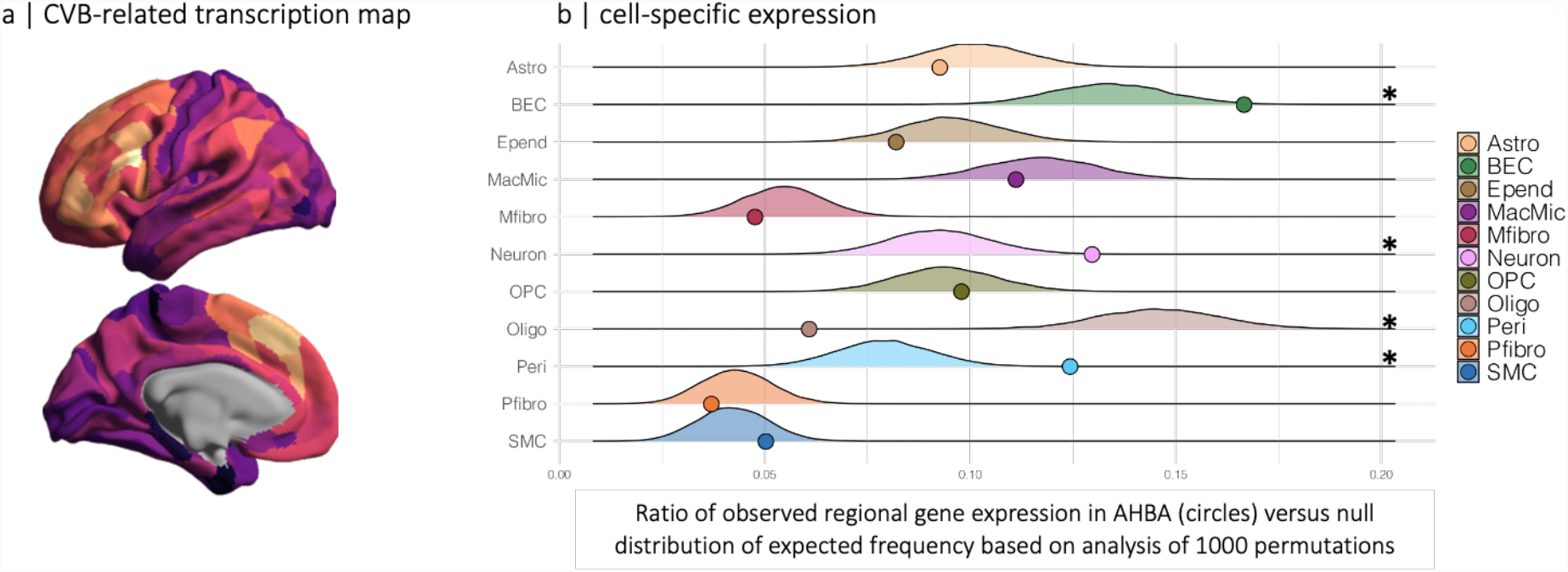
a, Spatial map in the transcriptome data related to the COVID-19-induced cerebrovascular burden map (CVB). b, Cell-type decomposition was used to identify cell-type enrichment based on extent to which genes expressed the transcriptome map in a. Gene sets for each cell-type was constructed by thresholding the top 70% of genes with greatest loadings. Note that results were consistent across a range of thresholds, ranging from 10% to no threshold, SI Figure 8. The ratio of genes in each gene set preferentially expressed in eleven distinct cell-types (circles) is shown against their null distribution of a model with random selection of all genes (10.000 permutations, *p-value < 0.05). For example, pericyte’s ratio is calculated from the number of genes preferentially expressed in pericytes divided by the total number of genes. Cell type-specificity of genes is described elsewhere (Yang et al., 2021) Astro – astrocytes, BEC – brain endothelial cells, Epend – ependymal, MacMic – macrophage/microglia, Mfibro – meningeal fibroblast, Neuron – neuron, OPC – oligodendrocyte precursor cells, Oligo – oligodendrocytes, Peri – pericytes, Pfibro – perivascular fibroblast, SMC – smooth muscle cells

## 4. Discussion

We show that abnormalities in cerebral microvascular function, measured using resting state fluctuation amplitudes (RSFA), persist many months after hospitalisation for acute COVID-19. The location of these abnormalities in lateral frontal and temporoparietal regions aligns partly with cerebrovascular dysfunction reported in association with ageing (Tsvetanov et al., 2020b, 2015), preclinical Alzheimer’s disease (Millar et al., 2020a) and systemic cardiovascular health (Tsvetanov et al., 2020b).These post-COVID-19 effects were observed over and above age. They are related to severity of the acute illness and the host response in the acute stage. These effects also relate to the post-COVID-19 cognitive function, common indices of mental health, and quality of life at an average of six months after hospitalisation.

In exploratory analysis, we found overlap between the regional distribution of this cerebrovascular impairment and spatial distribution of the vasoreactive receptor 5-HT1b and regions with high metabolic demands. 5-HT1b receptor is the dominant contractile 5-HT receptor in cerebral arteries (Barnes et al., 2021; Nilsson et al., 1999), acting on vasoconstriction by contracting smooth muscle endothelial smooth muscle directly or as moderator of other vasoconstrictors. Distinct from its effects on vascular tone, the presynaptic 5-HT1b receptor also has an important microvascular anti-inflammatory role, both in the cerebrovascular bed and more generally. Further, its loss has been implicated in progressive cognitive loss and abnormal modulation through descending serotoninergic outputs (Gharishvandi et al., 2020; Heijmans et al., 2021; Mitsikostas et al., 2002; Sibille et al., 2007), which may be relevant for late sequelae after COVID-19.

The overlap of cerebrovascular impairment with the regional distribution of aerobic glycolysis and glucose metabolism is spatially concordant with previous reports of hypometabolism in the subacute phase of COVID-19 (Hosp et al., 2021) and neurodegeneration (Vlassenko et al., 2010). This discussion provides a potential link between cerebrovascular impairment and metabolic dysfunction in frontoparietal regions, which could provide important insights regarding the mechanisms of late neurocognitive dysfunction following COVID-19 infection. Future work should establish whether changes to the microvasculature lead to hypometabolism (Shi et al., 2016) or the vulnerability of brain physiology in chronic phase is due to hypoxic and hypometabolic exposure in the subacute phase (Vestergaard et al., 2020). Collectively, these results demonstrate that the distribution of chronic cerebrovascular impairment related to COVID-19 severity maps to the spatial distribution of processes involved in coordinating metabolic and vasoregulatory responses associated with changes in brain function and cognition.

We also tested whether genes highly expressed in regions with COVID-19-induced cerebrovascular change are preferentially expressed in specific cell types using a molecular atlas of human brain vasculature (Yang et al., 2021). Dominant genes were overexpressed in perycites, brain endothelial cells and neurons, but underexpressed in oligodendrocytes. The evidence that genes implicated are enriched in pericytes and endothelial cells is particularly interesting. Brain endothelial cells are susceptible to direct SARS-CoV-2 infection through flow-dependent expression of ACE2. The SARS-CoV-2 S protein binding triggers a gene expression profile that may compromise the neurovascular interface (Kaneko et al., 2021). On the abluminal aspect of the neurovascular interface, pericytes express abundantly the angiotensin-converting enzyme-2 (ACE2) receptor (He et al., 2020). The expression can be increased with by exposure to the viral S protein, and importantly, potentiated in combination with hypoxia (Khaddaj-Mallat et al., 2021), a mechanism which could account for the modulation of RSFA abnormality by disease severity in our cohort.

Given this biological context the correlation of RSFA abnormalities with disease severity is open to two potential interpretations. One possibility is that these changes in cerebrovascular regulatory integrity are the consequence of direct viral invasion; while the other is that these abnormalities are a consequence of the inflammatory host response, which is a consequence of, but may not scale precisely with, viral infection. Spatial correlation of RSFA abnormality with the expression of ACE2 and Neuropilin-1, or of genes involved in cellular responses to viral invasion would have provided supportive evidence of a role for direct viral infection as a mechanism, but we were unable to demonstrate such a correlation. This negative finding favours the explanation that host inflammatory responses may be drivers in this context, and merit further investigation as mechanisms of late cerebrovascular regulatory dysfunction. It is important to acknowledge that our correlations with regional gene expression are based on expression patterns in normal brain, and that these expression patterns may be substantially altered by the inflammatory milieu that prevails in the context of COVID-19. Consequently, direct examination gene expression patterns in late COVID-19 survivors would provide additional insights.

Our study has several limitations. We are limited by the relatively small sample size, and by the absence of longitudinal imaging data. We also do not draw any causal inferences from the associations we observe. However, the demonstration of functional microvascular abnormalities following COVID-19 is important to understand the potential mechanisms of persistent cognitive and mental health problems. The association of microvascular abnormalities with late outcomes of relevance to patients, and the fact that they represent an easily accessible biomarker, suggest both a potential therapeutic target and/or a biomarker of treatment effect in interventional studies. It remains to be shown whether the localisation of RSFA abnormalities to regions rich in 5HT-1b receptors is a consequence of overactivity of these receptors (resulting in low cerebral blood flow), underactivity or loss of these receptors (resulting in vasoparalysis and/or inflammation), or a manifestation of flow-metabolism mismatching with inadequate substrate and oxygen delivery. This is relevant as potential therapeutic agents that are available to modulate both 5HT-1b function (Barnes et al 2021) and inflammatory response (Zhang et al., 2020) (Group et al., 2021b; Zhang et al., 2020).

In summary, we demonstrate that the severity of acute COVID-19 predicts cerebrovascular impairment six months later. The cerebrovascular abnormality was associated with worse cognitive function, mental health, functional recovery, and quality of life months after hospitalisation. Localised across lateral frontotemporoparietal regions, we show that its genetic signature shapes the composition of cell-types and metabolism. Collectively, these results implicate the COVID-19-related vulnerability of brain systems differentially relying on diverse physiology and cell biology in support of their functional specialisation.

## Data Availability

Code and composite data to reproduce manuscript figures, statistical analyses are available at https://github.com/kamentsvetanov/covid19_cerebrovascularburden.

## 5. Acknowledgements

This research was supported by the NIHR Cambridge Biomedical Research Centre (BRC-1215-20014), NIHR funding to the NIHR BioResource (RG94028 & RG85445), Guarantors of Brain (G101149), Wellcome Trust (220258), Medical Research Council (SUAG/051G101400; and SUAG/010 RG91365), and by the Addenbrookes Charitable Trust. We thank NIHR BioResource volunteers for their participation, and gratefully acknowledge NIHR BioResource centres, NHS Trusts and staff for their contribution. We thank the National Institute for Health Research, NHS Blood and Transplant, and Health Data Research UK as part of the Digital Innovation Hub Programme. The views expressed are those of the author(s) and not necessarily those of the NHS, the NIHR or the Department of Health and Social Care.

