## Supplemental Information for "Hospitalisation for COVID-19 predicts long lasting cerebrovascular impairment: A prospective observational cohort study"

Supplementary Information: Covid-19 severity and cerebrovascular burden

### 1. Supplementary Methods

#### 1.1. Imputations

Rate of missing inpatient, clinical and cognitive data varied between 0 and 38% (see Table 1); hence, to increase statistical power and efficiency, missing data were imputed before their analysis with neuroimaging. Incomplete variables were imputed under fully conditional specification, using the default settings of the multivariate imputation by chained equations (MICE) in R (Buuren and Groothuis-Oudshoorn, 2011). Multiple versions of each dataset were created (m = 5). Instead of accounting for variability in the parameter estimates between imputed datasets, we report any differences in the significance of parameters input to the multiple linear regression. For comparison, we also performed the analysis on the subset of complete cases.

#### 1.2. Data reduction

Data sets of interest stemmed from a range of modalities and different time-points (blood, cardiorespiratory, mental health and RSFA measures). To make these data sets tractable for univariate stages of the analytical approach (see below), we analysed a set of summary measures for each of the modality (also known as features or components) as illustrated in Figure 1. This had two advantages. First, it reduced the number of statistical comparisons. Second, it improved interpretability of the signals in the data by means of denoising and simplification given; low-dimensional data representations remove noise but retain signal of interest that can be instrumental in understanding hidden structures and patterns of multivariate data. Data reduction in the RSFA data can separate spatially overlapping sources of signal with different aetiologies (Xu et al., 2013), as it is known that cardiovascular *versus* cerebrovascular signals may vary across individuals and brain region in RSFA (Tsvetanov et al., 2020, 2015).

##### 1.2.1. Cerebrovascular components using RSFA data

We used independent component analysis (ICA) across participants to derive spatial patterns of RSFA maps across voxels. To this end, all RSFA maps were concatenated and submitted to Source-based Cerebrovasculometry (SBC) to decompose images across all individuals in a set of spatially independent sources without providing any information about the group (Xu et al., 2009), using the GIFT toolbox. Specifically, the n-by-m matrix of participants-by-voxels was decomposed into: (1) a source matrix that maps each independent component to voxels (referred to as $RSFA_{IC}$ maps), (2) a mixing matrix that maps $RSFA_{IC}$ to participants i.e. subject scores (1 per participant) indicating the degree to which a participant expresses a defined $RSFA_{IC}$. Subject scores were used in subsequent analyses to understand their relationship with other variables of interest as described in section 'Analytical approach' below.

To confirm the generalisation of the results across analyses and datasets we performed a second-level univariate analysis in SPM12 with RSFA as dependent variable. Group identity, age and sex were defined as predictors. The spatial map of the group effects was correlated with the spatial map of the independent component differentiating patients and controls. Spatial maps of age and sex effects were compared with those reported in previous reports on large-scale population-based samples (n = 226, Tsvetanov et al., 2020). Visualisation and correlation across maps were carried out on maps thresholded at uncorrected p-values of 0.05 for more complete description of the spatial representation.

##### 1.2.2. Covid-19 Severity component using inpatient data

We used principal component analysis to derive composite variables within each of the three non-imaging datasets – Covid-19 severity dataset during inpatient care, cardiorespiratory dataset during clinic visit and cognitive and mental health dataset during research visit. The nine measures constructing the Covid-19 severity component (Covid-19 $Severity_{PC1}$) included Covid-19 WHO Progression Scale and blood markers (CRP, ferritin, IL-6, bilirubin, D-dimer, PT, APTT and iPlatelets). All variables were normalised to a mean of 0 and standard deviation of 1. C-reactive protein (CRP) and bilirubin were assayed on the Siemens Advia 2400; interleukin-6 and ferritin on the Siemens Centaur; platelet count on the Siemens ADVIA 2120; and prothrombin time, activated partial thromboplastin time, and D-dimer on the Siemens ACL-TOP (all devices manufactured by Siemens Healthcare GmbH, Erlangen, Germany).

##### 1.2.3. Cardiorespiratory dysfunction components using clinical visit data

Cardiorespiratory dysfunction was represented by the first two principal components ($CRD_{PC1}$ and $CRD_{PC2}$) constructed from blood pressure (lying and standing pulse pressure, and orthostatic systolic and diastolic blood pressure), differences in heart rate and blood oxygenation during a 6-minute walk test, and spirometry function (first principal component from spirometry data). All variables were normalised to a mean of 0 and standard deviation of 1.

##### 1.2.4. Quality of life, cognition and mental health components using research visit data

Quality of life, cognition and mental health were represented by the first two components from MoCA, imRS, BI, iGAD-7, iPCL-5, iPHQ-9, iPHQ-15 total scores, and eight SF-36 subscores. All variables were normalised to a mean of 0 and standard deviation of 1.

### 2. Supplementary Figures and Analyses

#### 2.1. Analytical strategy

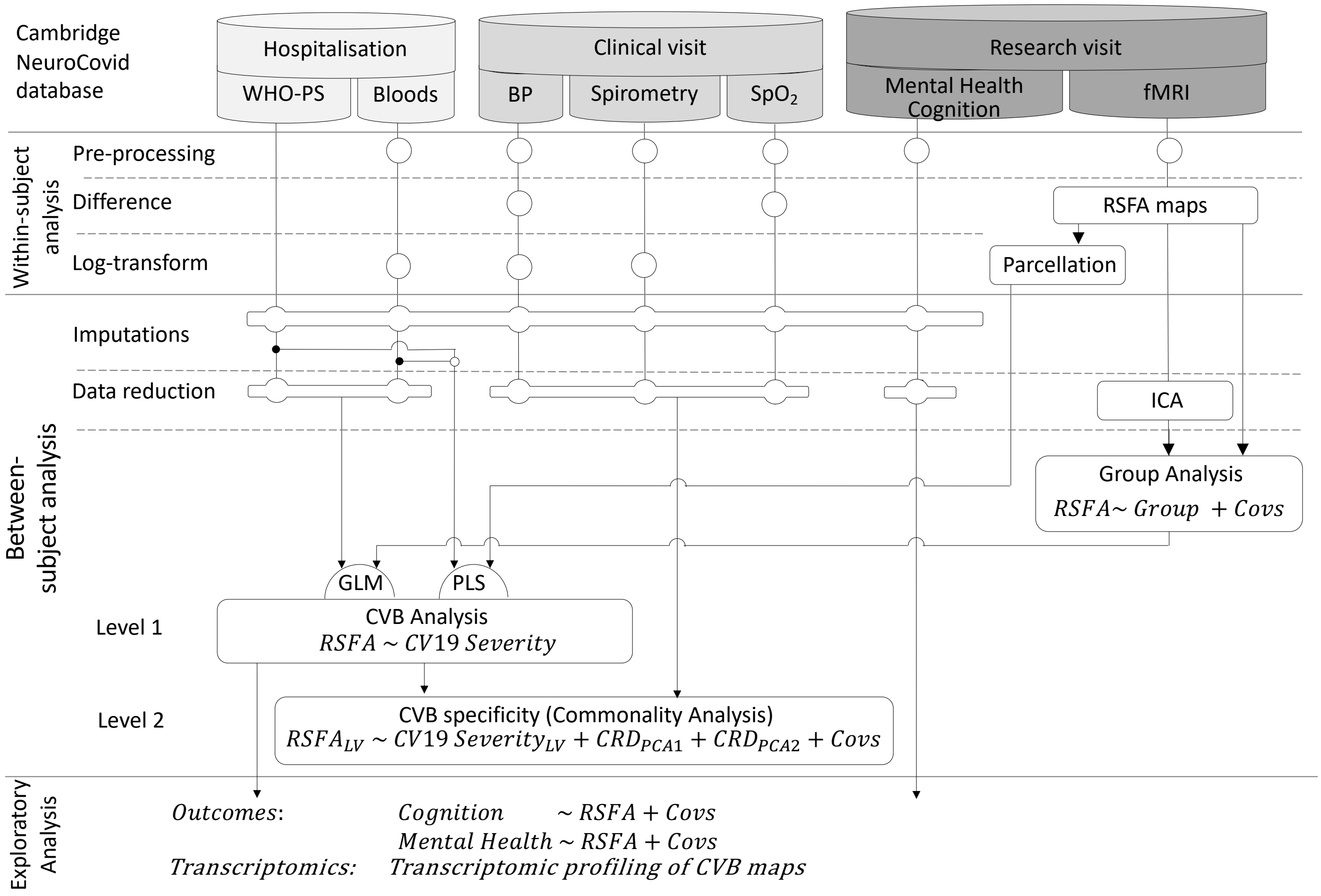


*SI Figure 1.* Schematic representation of various modality datasets in the study, their processing pipelines on a within-subject level, as well as data-reduction techniques and analytical strategy on between-subject level to test for associations between acute Covid-19 Severity and chronic cerebrovascular impairment. WHO-PS, Covid-19 WHO progression scale; BP, blood pressure; SpO2, blood oxygen saturation; fMRI, functional magnetic resonance imaging; RSFA, resting state fluctuation amplitudes; PCA, principal component analysis; ICA, independent component analysis; Covs, covariates of no interest; GLM, general linear model; PLS, partial least squares; LV, latent variable from PLS analysis; CRD, cardiorespiratory dysfunction component; CVB, cerebrovascular burden component;

#### 3.1. Imputations

The percentage of missing values across the inpatient, clinical visit and research visit variables varied between 0 and 38%. In total 69 out of 405 records (17%) were incomplete for impatient data; 71 out of 360 records (20%) were incomplete for clinical visit data; and 121 out of 675 records (18%) were incomplete for cognitive and mental health data.

#### 3.2. Data reduction

##### 3.2.1. Covid-19 Severity component using inpatient data

A single component of Covid-19 Severity was informed using principal component analysis on inpatient data including Covid-19 WHO Progression scale and eight biomarkers previously associated with severity in patients hospitalised with Covid-19. The component explained 40% of the total variance loaded most strongly on Covid-19 WHO Progression scale, C-Reactive Protein, D-Dimer and serum ferritin, followed APTT and bilirubin. Other biomarkers loaded in the expected direction, but to a lesser extent.

##### 3.2.2. Cardiorespiratory dysfunction

To balance the representativeness of each data type in the imputations and data-reduction stages, spirometry measures were reduced to a single  variable using principal components analyses. In total, 12 spirometry measures, based on three sets of FEV1, FVS, PEF and FEV1/PEV, were submitted to PCA. The first component expressed FEV1 and FVC values explaining 45% in the data.

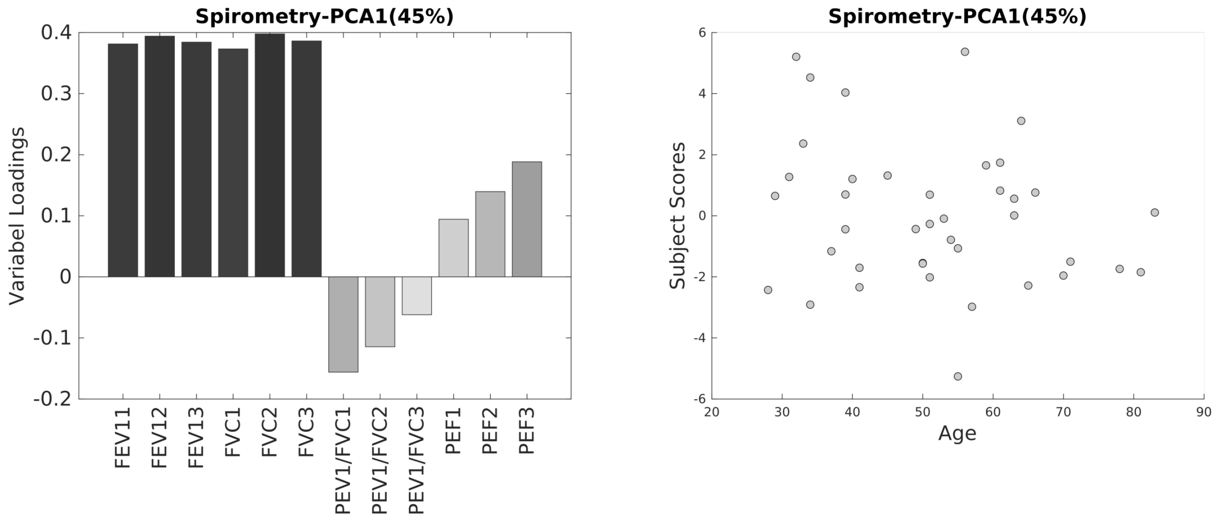


*SI Figure 2. Variable loading and subject scores for the first principal component on spirometry data.*

Cardiorespiratory dysfunction was represented by the first two principal components ($CRD_{PC1}$ and $CRD_{PC2}$) constructed from blood pressure (lying and standing pulse pressure, and orthostatic systolic and diastolic blood pressure), differences in heart rate and blood oxygenation during a 6-minute walk test, and spirometry function (first principal component from spirometry data). The first compoenent explained 37% loading highly on lung function, oxygen saturation and pulse pressure. The second component, explaining 22%, loaded on oxygen saturation and orthostatic hypotension.

##### 3.2.3. Mental health and cognition

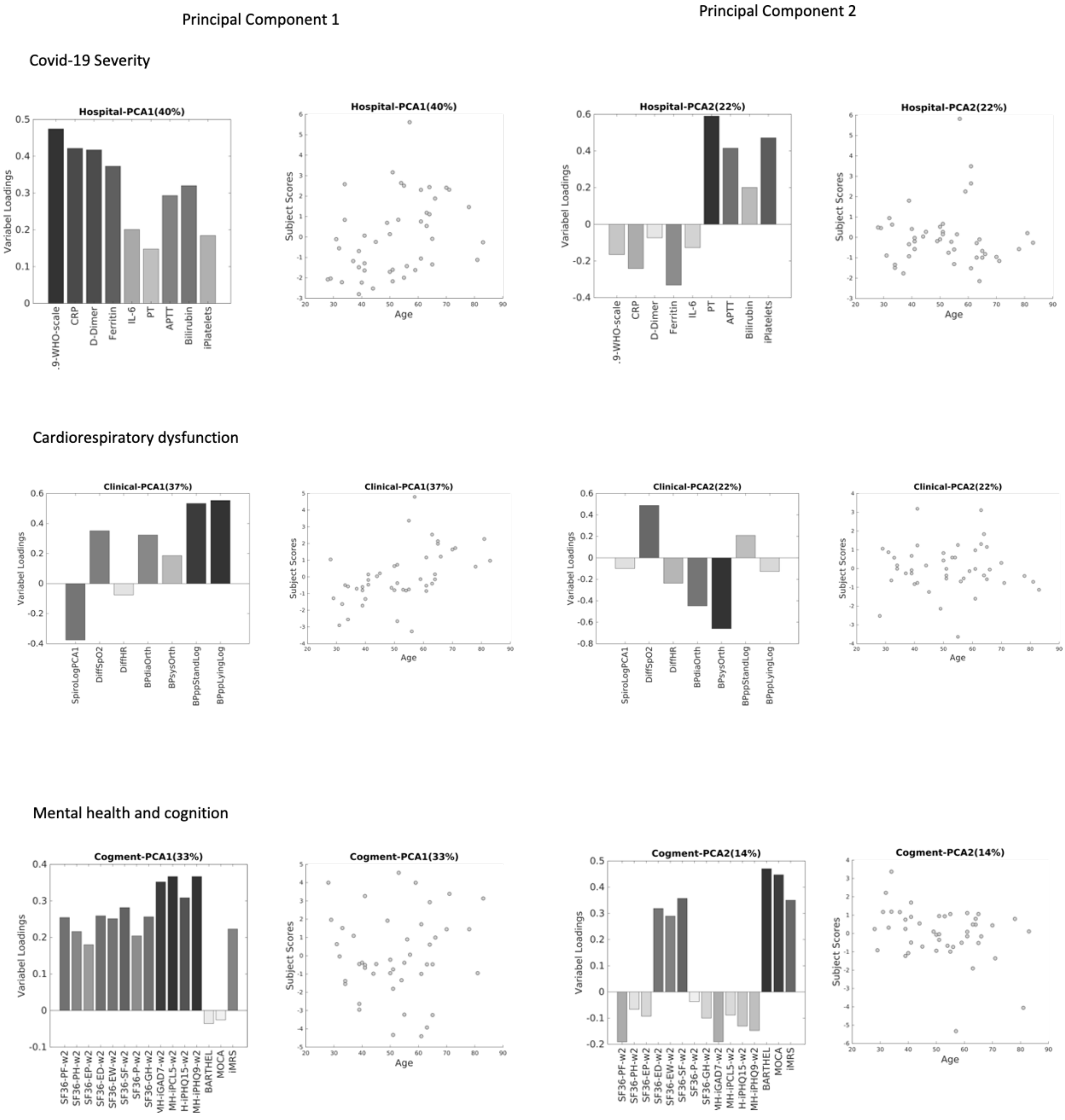


*SI Figure 3. Variable loading and subject scores for the first two principal component analysis on Covid-19 severity data (row 1), cardiorespiratory dysfunction (row 2) and mental health and cognition (row 3).*

##### 3.2.4.Cerebrovascular burden in RSFA data

To characterise cerebrovascular components in the entire dataset (42 controls and 45 patients) without providing group information, we used independent component analysis to determine a small set of spatially independent components in the RSFA data (n=8) as informed by minimum distance length criteria. Spatial maps for all components is shown in SI Figure 4.

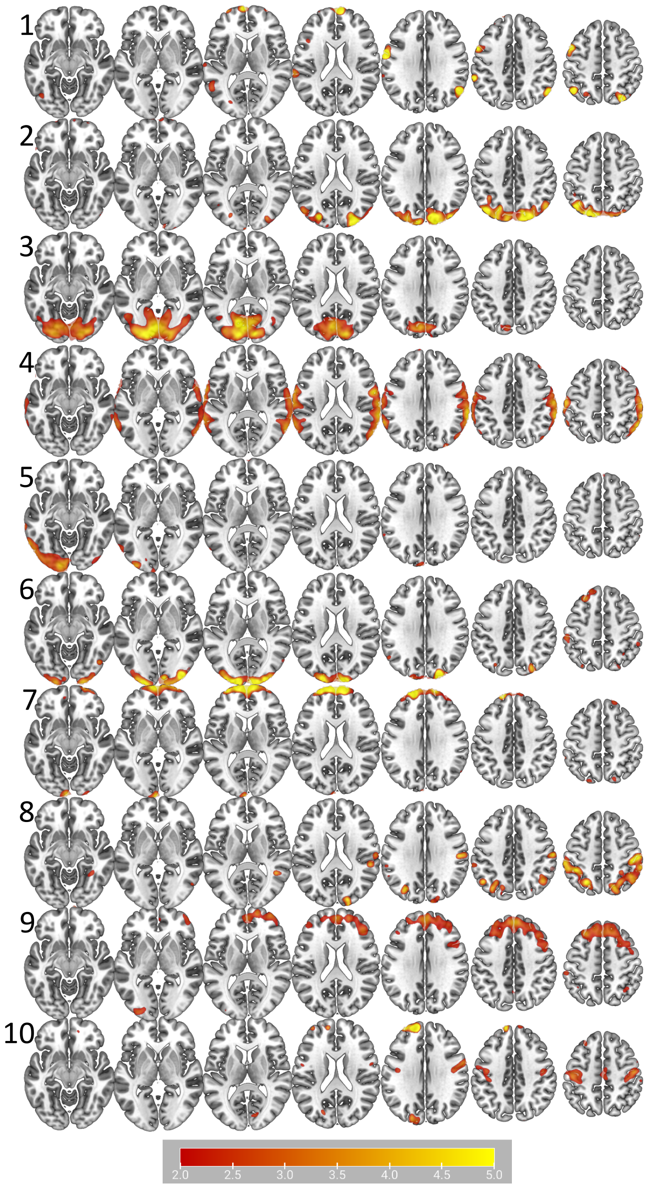


*SI Figure 4. Indepenendent component analysis maps for all 10 components. Maps are thresholded at T-value >2 for more complete description of the spatial representation.*

The spatial pattern of the component differentially expressed between patients and controls, (IC4), was consistent with the spatial pattern from a univariate analysis.

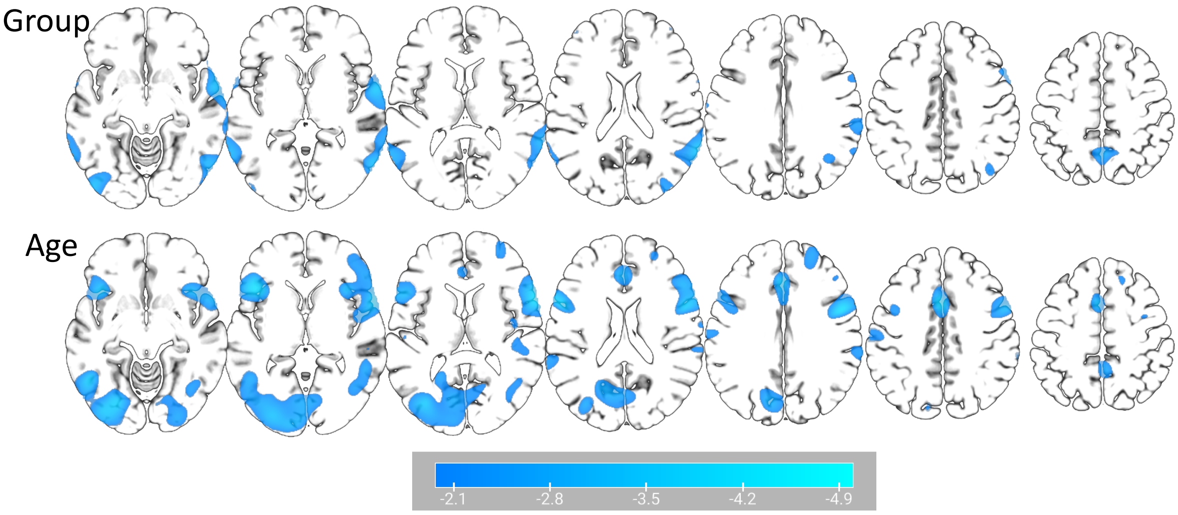


*SI Figure 5. Voxel-wise association between RSFA and group identity (patient vs control) and age. The spatial pattern of group effects was highly consistent with the pattern identified using ICA (IC4 in SI Figure 3), r=.46, p-spin<.001. The spatial pattern associated with age effect was highly consistent with the pattern derived from a previous study using a large population-based cohort* (n=226, Tsvetanov et al., 2020), r=.42, p-spin<.001. *Maps are thresholded at uncorrected p-values of 0.05 for more complete description of the spatial representation.*

#### 3.3. Relationship between Covid-19 Severity Component and voxel-wise RSFA

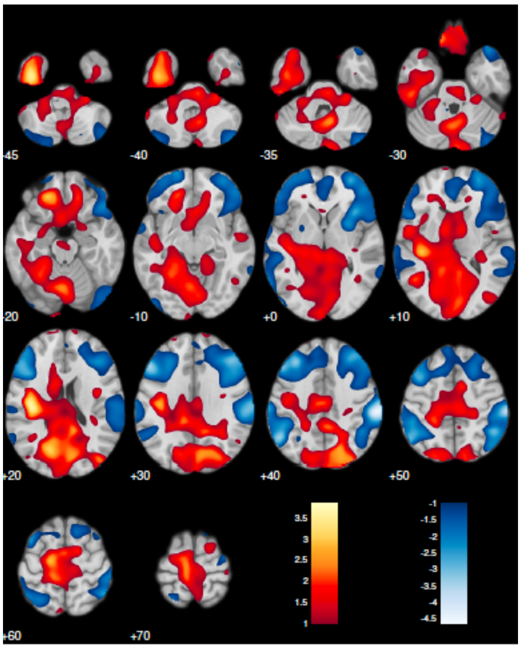


*SI Figure 6. Association between Covid-19 Severity PCA1 and RSFA on voxel level showing a negative association between Covid-19 Severity and RSFA values in frontal and temporoparietal regions.*

#### 3.4. Multivariate relationship between Covid-19 Severity and RSFA

*SI Table 1. Association between cerebrovascular impairment and Covid-19 Severity identified by partial least squares analysis cannot be explained by covariates including age, sex and cardiorespiratory dysfunction.*

```
Linear regression model (robust fit):
    RSFA ~ 1 + CV19Severity + Age + Sex + Clinical_PCA1_iAll + Clinical_PCA2_iAll

Estimated Coefficients:
                          Estimate        SE        tStat       pValue
                         _________    _______    ________    _________

    (Intercept)          -0.064952    0.11765    -0.55208      0.58404
    CV19Severity           0.46836    0.13855      3.3803    0.0016562
    Age                    0.23067    0.14447      1.5967       0.1184
    Sex                   0.063285    0.12431     0.50908      0.61357
    Clinical_PCA1_iAll   0.0063342    0.14495      0.0437      0.96537
    Clinical_PCA2_iAll     0.13683    0.11962      1.1439      0.25963

Number of observations: 45, Error degrees of freedom: 39
Root Mean Squared Error: 0.789
R-squared: 0.406,  Adjusted R-Squared: 0.329
F-statistic vs. constant model: 5.32, p-value = 0.000801
```

*SI Table 2. Results from commonality analysis using 10.000 permuations indicating that a portion of the variance between Covid-19 Seveirty-LV and RSFA-LV was explained by age (PercentTotal for 'CV19Severity,Age'), and shared variance between age and cardiorepriratory dysfunction component 1 (CV19Sevitiy,Age,Clinical_PCA1_iAll). Covid-19 Severity remained as the largest unique predictor of RSFA-LV (CV19Severity).*

| | Coefficient | PercentTotal | tR2 | pPerm |
|---|---|---|---|---|
| CV19Severity | 0.16967 | 0.37385 | 2.823 | 0.0062 |
| Age | 0.070974 | 0.15639 | 1.7261 | 0.0788 |
| Sex | 0.013894 | 0.030615 | 0.74129 | 0.4386 |
| Clinical_PCA1_iAll | 0.00023696 | 0.00052212 | 0.096144 | 0.9164 |
| Clinical_PCA2_iAll | 0.030467 | 0.067133 | 1.1071 | 0.2426 |
| CV19Severity,Age | 0.05626 | 0.12396 | 1.5248 | 0.0002 |
| CV19Severity,Sex | -0.012359 | -0.027232 | -0.6986 | 0.062 |
| Age,Sex | -0.0074316 | -0.016375 | -0.54037 | 0.0922 |
| CV19Severity,Clinical_PCA1_iAll | 0.011228 | 0.024741 | 0.66549 | 0.1278 |
| Age,Clinical_PCA1_iAll | 0.0109 | 0.024017 | 0.65558 | 0.1902 |
| Sex,Clinical_PCA1_iAll | -0.00016011 | -0.00035279 | -0.079027 | 0.4254 |
| CV19Severity,Clinical_PCA2_iAll | 0.015614 | 0.034404 | 0.78651 | 0.0032 |
| Age,Clinical_PCA2_iAll | -0.0047483 | -0.010463 | -0.43135 | 0.035 |
| Sex,Clinical_PCA2_iAll | -0.00024515 | -0.00054017 | -0.097792 | 0.1198 |
| Clinical_PCA1_iAll,Clinical_PCA2_iAll | 6.6492e-05 | 0.00014651 | 0.050925 | 0.3616 |
| CV19Severity,Age,Sex | 0.0067462 | 0.014865 | 0.51468 | 0.0302 |
| CV19Severity,Age,Clinical_PCA1_iAll | 0.089032 | 0.19618 | 1.9523 | 0 |
| CV19Severity,Sex,Clinical_PCA1_iAll | 0.00017001 | 0.00037461 | 0.081435 | 0.4196 |
| Age,Sex,Clinical_PCA1_iAll | 8.9853e-05 | 0.00019799 | 0.0592 | 0.453 |
| CV19Severity,Age,Clinical_PCA2_iAll | -0.00052556 | -0.001158 | -0.14321 | 0.1966 |
| CV19Severity,Sex,Clinical_PCA2_iAll | -0.0001444 | -0.00031818 | -0.07505 | 0.3434 |
| Age,Sex,Clinical_PCA2_iAll | 0.00029183 | 0.00064304 | 0.1067 | 0.132 |
| CV19Severity,Clinical_PCA1_iAll,Clinical_PCA2_iAll | 0.00070192 | 0.0015466 | 0.16551 | 0.1896 |
| Age,Clinical_PCA1_iAll,Clinical_PCA2_iAll | -0.0012918 | -0.0028464 | -0.2246 | 0.0962 |
| Sex,Clinical_PCA1_iAll,Clinical_PCA2_iAll | -2.57e-05 | -5.6629e-05 | -0.03166 | 0.1456 |
| CV19Severity,Age,Sex,Clinical_PCA1_iAll | 0.0035495 | 0.0078211 | 0.37272 | 0.0986 |
| CV19Severity,Age,Sex,Clinical_PCA2_iAll | 0.00032904 | 0.00072502 | 0.1133 | 0.0456 |
| CV19Severity,Age,Clinical_PCA1_iAll,Clinical_PCA2_iAll | 0.00024475 | 0.00053929 | 0.097712 | 0.285 |
| CV19Severity,Sex,Clinical_PCA1_iAll,Clinical_PCA2_iAll | 2.1576e-05 | 4.7541e-05 | 0.029008 | 0.2166 |
| Age,Sex,Clinical_PCA1_iAll,Clinical_PCA2_iAll | 2.8302e-05 | 6.2361e-05 | 0.033224 | 0.159 |
| CV19Severity,Age,Sex,Clinical_PCA1_iAll,Clinical_PCA2_iAll | 0.00025444 | 0.00056065 | 0.099629 | 0.0062 |

#### 3.5. Relationships between RSFA and Cognitive and Mental health

*SI Table 3. Association between* physical, cognitive, and mental functioning *(PCM) component and Covid-19-related cerebrovascular impairment where age and sex are included as covariates of no interest.*

```
Linear regression model (robust fit):
    Cogment_PCA2_iAll ~ 1 + RSFA + Age + Sex

Estimated Coefficients:
                     Estimate         SE         tStat       pValue
                   __________    _______    _________    ________

    (Intercept)    -0.0095463    0.11893    -0.080267     0.93642
    RSFA             -0.35209     0.1367      -2.5757    0.013708
    Age              -0.14281    0.13975      -1.0218     0.31285
    Sex              -0.29709    0.12336      -2.4082    0.020612

Number of observations: 45, Error degrees of freedom: 41
Root Mean Squared Error: 0.798
R-squared: 0.294,  Adjusted R-Squared: 0.242
F-statistic vs. constant model: 5.69, p-value = 0.00236
```

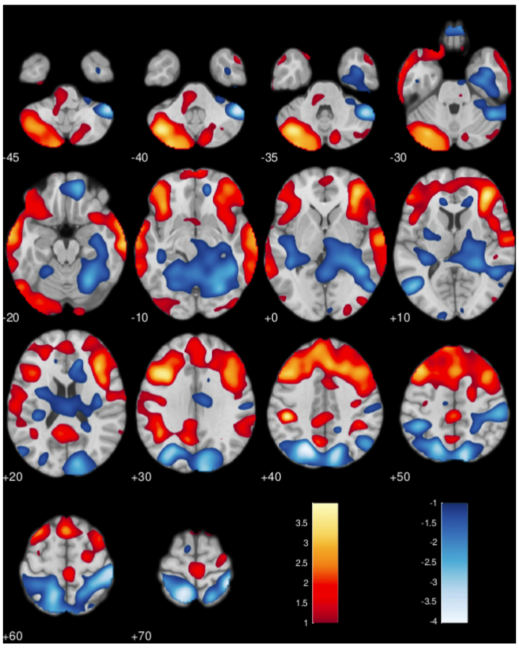


*SI Figure 7 Association between* physical, cognitive, and mental functioning *(PCM) component and RSFA on voxel level showing a positive association between phsycial and cognitive functioning with RSFA values in frontal and temporall regions.*

a | CVB-related transcription map b | cell-specific expression

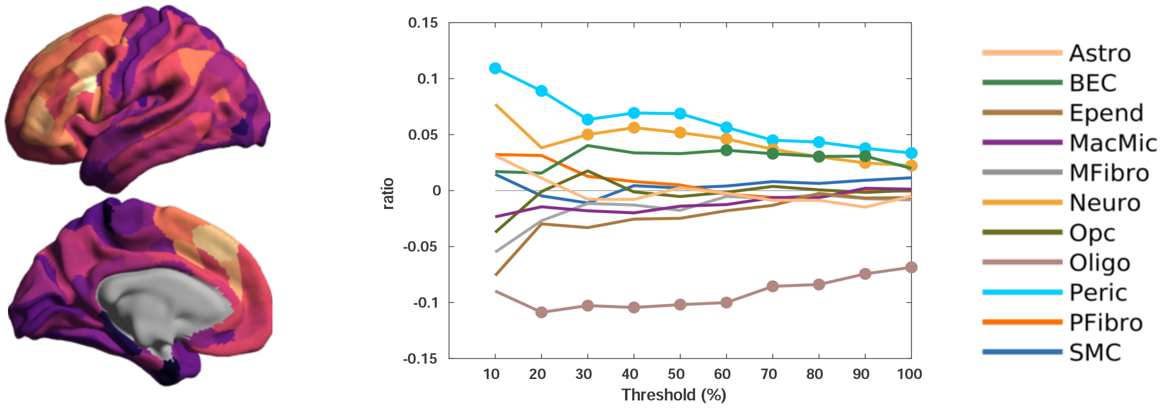


*SI Figure 8. Specific cell-type expression in Covid-19 induced cerebrovascular burden constructed from the largest positive loadings, ranging from the top 10% genes to all genes. Each point represents the difference between the ratio of genes in each gene set preferentially expressed in a cell-type and the mean null ratio, computed from null distribution of random gene sets (10.000 permutations),* (Hansen et al., 2021)*. Curves above zero indicate overexpression and curves below zero indicate underexpression. Circles demonstrate significance. Astro – astrocytes, BEC – brain endothelial cells, Epend – ependymal, MacMic – macrophage/microglia, MFibro – meningeal fibroblast, Neuro – neuron, Opc – oligodendrocyte precursors, Oligo – oligodendrocytes, Peri – pericytes, PFibro – perivascular fibrblast, SMC – smooth muscle cells.*
